# Transfer Learning for COVID-19 Pneumonia Detection and Classification in Chest X-ray Images

**DOI:** 10.1101/2020.12.14.20248158

**Authors:** Iason Katsamenis, Eftychios Protopapadakis, Athanasios Voulodimos, Anastasios Doulamis, Nikolaos Doulamis

## Abstract

We introduce a deep learning framework that can detect COVID-19 pneumonia in thoracic radiographs, as well as differentiate it from bacterial pneumonia infection. Deep classification models, such as convolutional neural networks (CNNs), require large-scale datasets in order to be trained and perform properly. Since the number of X-ray samples related to COVID-19 is limited, transfer learning (TL) appears as the go-to method to alleviate the demand for training data and develop accurate automated diagnosis models. In this context, networks are able to gain knowledge from pretrained networks on large-scale image datasets or alternative data-rich sources (i.e. bacterial and viral pneumonia radiographs). The experimental results indicate that the TL approach outperforms the performance obtained without TL, for the COVID-19 classification task in chest X-ray images.

## 1 INTRODUCTION

The first human cases of COVID-19 were reported in Wuhan, China, in December 2019. Given its rapid spread, a month later, the World Health Organization (WHO) declared novel coronavirus (2019-nCoV) a public health emergency of international concern [1] and a pandemic on 11 March 2020 [2]. The COVID-19 pandemic, as of September 27, 2020, has resulted in over 33 million confirmed cases and over 1 million deaths worldwide [3], causing significant socio-economic repercussions on people’s livelihoods [4]. Just for the sake of comparison, the SARS outbreak in 2002-2003 caused 774 deaths in 26 countries, with a total of 8,098 infected people [5], [6].

Although the COVID-19 disease can cause multi-organ damages [7], the lungs are the most affected organs of the human body, due to their abundance of angiotensin-converting enzyme 2 (ACE2), which is a receptor that acts as the door, through which the virus is able to enter the host cells [8]. Therefore, the lungs are the focal point of the human body, for the COVID-19 implications detection.

On this basis, the role of chest radiography in the diagnosis of COVID-19 disease is crucial. In particular, like other types of pneumonia (e.g. bacterial pneumonia), on thoracic X-rays of patients with COVID-19 pneumonia, is observed an increased (whiter) density of lungs. Usually, the more severe the disease, the more intense the whiteness on the chest X-rays. In the typical case, ground-glass opacity occurs, with possible simultaneous existence of linear opacities (e.g. peripheral horizontal white lines), resulting in slightly obscured lung markings (Figure 1a). It is underlined that in severe cases the lung markings become invisible due to the dense whiteness, a phenomenon known as consolidation (Figure 1b) [9]. Hence, utilizing chest radiography, or combining it with laboratory and clinical assessment, can be an efficient tool for the early and accurate diagnosis of COVID-19 pneumonia.

**Figure 1:**
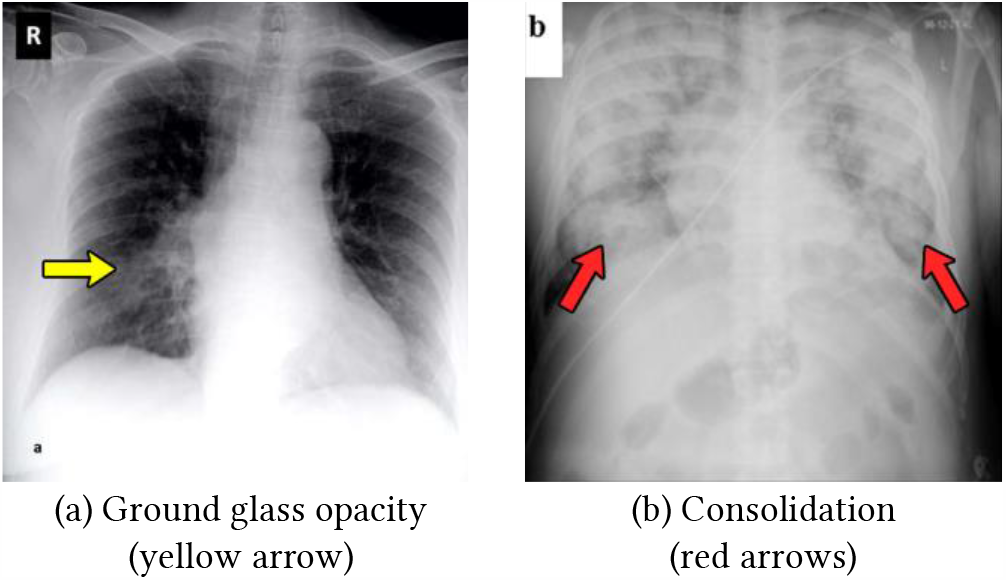
Increased whiteness in the lungs, because of COVID-19 pneumonia. [10].

During the last few years, the use of deep learning applications in medical imaging analysis has received much attention from the scientific community, surpassing in many cases the performance of medical professionals [11]. The pneumonia detection task on chest radiography can be interpreted as a computer vision classification problem, where the input is a thoracic X-ray image and the output indicates the existence or the absence of the infection. In this context, several studies have proven that convolutional neural networks (CNNs) show great performance on various image classification tasks, such as the one investigated in this paper [12], [13].

However, the main problem that arises in such techniques is the requirement for large-scale datasets in order to train and evaluate the classifiers [14]–[16]. At the time of writing this paper, the number of X-ray samples related to COVID-19 is extremely limited, hence transfer learning is a preferable method in order to train efficiently the deep CNNs [17]. More specifically, with transfer learning, we are able to avoid the training of the deep models from scratch and also the lack of training data, by taking advantage of the extraction of knowledge from other alternative tasks.

The purpose of this paper is to demonstrate the efficacy of the state-of-the-art deep CNNs, trained through transfer learning, in giving accurate pneumonia diagnosis from frontal-view chest X-rays. The proposed models will be evaluated according to their performance in detecting traces of COVID-19 infection in the patients’ thoracic X-ray images, in conjunction with their ability to differentiate the specific patients from those infected with bacterial pneumonia.

The remainder of this paper is organized as follows. Section 2 briefly presents related work on applications of deep learning frameworks in medical imaging. Section 3 outlines the proposed computer vision methods for pneumonia classification, and Section 4 discusses the experimental results obtained from the deep models. Finally, Section 5 concludes the paper with a summary of findings.

## 2 RELATED WORK

Recent developments in deep learning applications, availability of large datasets and advances in the computing power have led algorithms to be a powerful auxiliary tool in the hands of medical professionals. Several studies have been carried out, emphasizing the high performance of deep models in a wide variety of medical imaging applications, such as, for instance, detection of diabetic retinopathy [18], identification of arrhythmia [19], early prognostication of Alzheimer’s disease dementia [20], diagnosis of brain hemorrhage [21], and classification of various types of cancer (e.g. breast [22], skin [23], brain [24], and prostate [25]).

Automated diagnosis of various chest diseases using radiographs has been gaining momentum, due to the latest breakthrough of artificial intelligence (AI) systems and the dramatic increase in available public thoracic X-ray datasets [26]. In detail, deep learning algorithms are increasingly set to become a key factor in pediatric pneumonia diagnosis [27] and classification of pulmonary tuberculosis [28]. Moreover, in the work of [29], the authors developed a deep model, based on the DenseNet-121 architecture, which is able to accurately detect the presence of 14 different thoracic pathologies, using chest radiographs. It is noted that their analysis has found that the deep learning algorithm has reached or even surpassed the performance level of clinical assessment by human radiologists, on 11 out of the 14 pathologies.

### 2.1 Our Contribution

Inspired by the above research work, in this paper, we introduce a deep learning framework that can detect COVID-19 pneumonia in thoracic X-rays. Considering the pandemic outbreak, the proposed framework aims to fill the gap between the limited number of highly skilled radiologists and the growing need for chest X-ray interpretation.

## 3 PROBLEM FORMULATION AND PROPOSED METHODOLOGY

### 3.1 Problem Formulation

Automated pneumonia diagnosis, based on chest X-ray images, is a computer vision problem that can be addressed as a (i) classification [11], [13], [35], (ii) object detection [36], [37], or (iii) semantic segmentation task [38], [39]. In the first procedure, the output of the deep learning model indicates the existence or the absence of the infection, in a given X-ray image. In the second method, the algorithm provides bounding boxes that indicate the location and the dimensions of the infected areas, in the patient’s lung. The latter approach involves pixel-based classification methods (e.g. Fully Convolutional Networks (FCN) [40], U-Nets [41], and Mask R-CNN [42]), thus defining precisely the shape of the symptomatic areas.

In this paper, we address the problem of pneumonia detection as a multi-class classification task. The input *X* ∈ ℝ^*w*×*h*×3^ of the problem is an RGB frontal-view thoracic X-ray image, with dimensions *w* × *h*, and the output *y* ∈ {0, 1, 2} denotes respectively the absence of pneumonia, the bacterial pneumonia infection, or the COVID-19 pneumonia infection.

### 3.2 Dataset Description

In this paper, we are using two different datasets, consisted of exclusively frontal-view chest X-ray images of various resolutions, ranging from 220×206 to 5623×4757.

The first dataset, referred as Dataset_0, comes from the work of Kermnay et al. [30] and includes 1,575 X-ray images of healthy patients (Figure 2a), 2,772 images of patients with bacterial pneumonia (Figure 2b) and 1,493 images of patients with viral pneumonia (Figure 2c). Dataset_0 was used for training a multi-class classification model, which will be targeting three classes: (i) normal healthy patients, (ii) patients with pneumonia caused by bacteria, (iii) patients with pneumonia caused by a virus.

**Figure 2:**
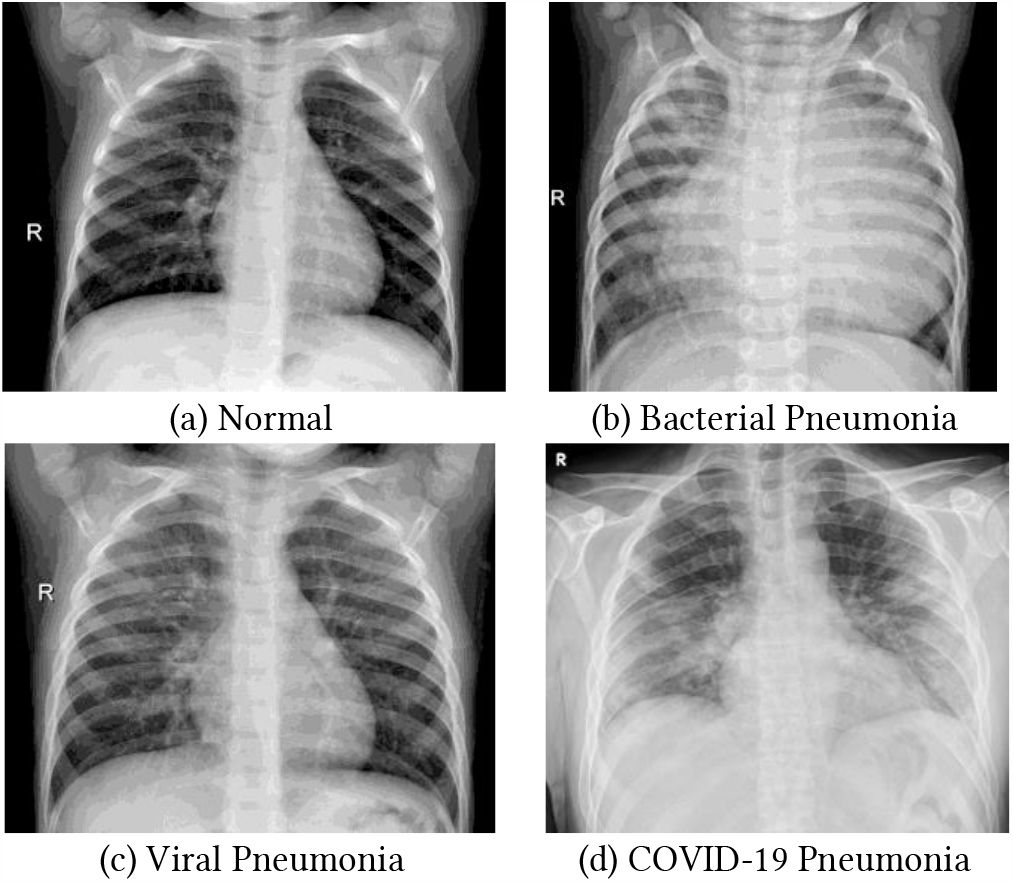
Examples of frontal-view chest X-Ray images from the datasets.

The second dataset, referred as Dataset_1, consists of thoracic X-ray images of confirmed COVID-19 cases (Figure 2d) that come from the COVID-19 image data collection of Cohen et al. [10], as well as a subset of the aforementioned Dataset_0. More specifically, Dataset_1 contains 544 images of healthy patients, 500 images of patients with bacterial pneumonia and 456 images of patients with COVID-19. It is underlined that through the undersampling process of the normal and bacterial classes, we can avoid imbalanced data problems, since at the time of writing this paper the number of X-ray samples for the COVID-19 class is extremely limited. From the whole number of chest X-ray images in Dataset_1, 80% is used for training the deep learning multi-class classification models, while the rest 20% is used for testing. Among the training data, 87.5% of them were used for training and the remaining 12.5% for validation.

### 3.3 Transfer Learning

The idea of transfer learning lies in the fact that, in machine learning, we can utilize knowledge gained from one problem A and then apply it to another related problem B [31]. For instance, knowledge obtained while learning to detect cats can be transferred when trying to detect dogs. It is emphasized that, generally speaking, transfer learning is more effective when tasks A and B are relatively correlated, since then, they possess similar low-level image features (e.g. edges, lines, corners, and curves) [32].

Furthermore, through transfer learning, we are able to create accurate deep models on small datasets, by leveraging existing networks, already trained on large datasets. Thus, instead of training our network from scratch, we can utilize these pretrained models, the low-level features of which are pretty generic, and therefore can be used in any image classification task [14], [33].

Based on the above and given the limited data of COVID-19 X-ray images, transfer learning is the preferred method, in order to develop accurate automated diagnosis models [17]. In this way, we are able to overcome the isolated learning paradigm of COVID-19 and utilize knowledge from other data-rich sources, such as the bacterial and viral pneumonia radiographs. Additionally, we leverage pretrained networks on large-scale image datasets, such as the ResNet-50 trained on ImageNet [34], and transfer their generic knowledge into our pneumonia classification problem.

### 3.4 Employed Network Architectures

In order to detect pneumonia cases from chest X-ray images, we designed a simple CNN, the architecture of which is depicted in Figure 3. Initially, a CNN, referred as Model_0 and based on the specific architecture, was trained on Dataset_0. Model_0 is able to classify the following three classes: (i) normal, (ii) bacterial pneumonia, and (iii) viral pneumonia. Then, two CNNs, based on the same architecture, were trained on Dataset_1: the first model, referred as Model_1, was trained from scratch using random initial weights, whereas the second model, referred as Model_2, was trained using transfer learning from Model_0. Finally, a ResNet-50 network, referred as Model_3, was also trained on Dataset_1, initialized with weights pretrained on ImageNet [34]. It is noted that the last fully connected layer of the ResNet-50 is replaced by one that can identify three classes instead of 1000. The last three models are able to classify the following three classes: (i) normal, (ii) bacterial pneumonia, and (iii) COVID-19 pneumonia.

**Figure 3:**
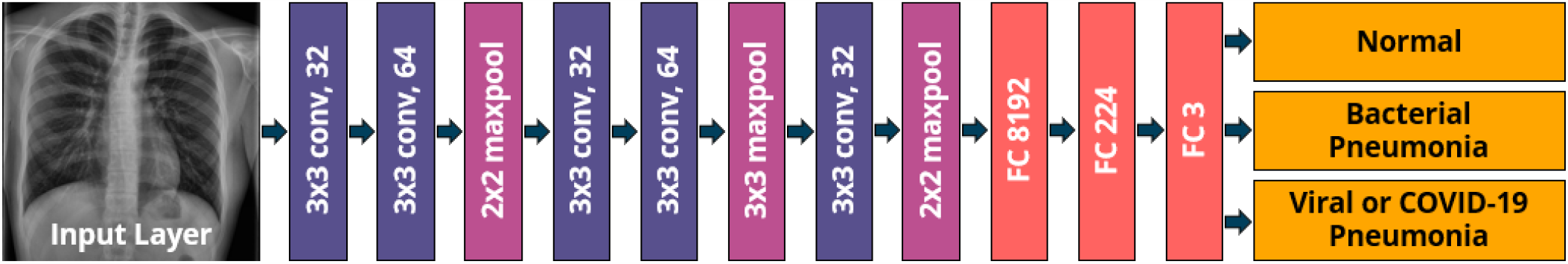
Architecture of the proposed CNN for pneumonia classification.

## 4 EXPERIMENTAL EVALUATION

### 4.1 Experimental Setup – Models Training

All the deep models were developed in Python, using Keras and TensorFlow libraries. The networks were trained and evaluated using an NVIDIA Tesla K80 GPU, provided by Google Colab.

We trained the CNNs, using batches of size 20, for 500 epochs, with early stopping criteria set to 10 epochs, to avoid overfitting. During data preprocessing, we resized input images to a resolution of 224 × 224 pixels.

All the models were trained using Adam optimizer with the default parameters (*β*_1_ = 0.9 and *β*_2_ = 0.999), with an initial learning rate of 0.001 that is reduced by a factor of 10 when there is no improvement in the validation loss, for five consecutive epochs. Regarding Model_2 and Molel_3, to which were applied transfer learning techniques, initially we tuned only the weights of the final dense layer and froze the rest of the networks. Then, we unfroze the whole Model_2 and the last 11 layers of Model_3 and continued the training procedure in order to fine-tune the reused layers. It is highlighted that after unfreezing the reused layers, the learning rate was initially set to 0.0001, in order to prevent damaging the reused weights. Lastly, during the models’ training process, we optimize the categorical cross-entropy loss

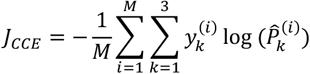

where M is the number of training examples, 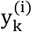 is the target probability that the i^th^ training instance belongs to the k^th^ class, and 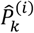 is the scalar value in the models’ outputs.

### 4.2 Results

To test the proposed approaches, we consider four performance metrics, to evaluate the results of the deep models on the test set of Dataset_1: (i) prediction, which measures the correctness of positive predictions (ii) recall, which refers to the ratio of total relevant instances correctly classified by the network, (iii) accuracy, which shows the proportion of correct predictions to the total predictions made by the classifier, and (iv) F1-score, which is the harmonic mean of precision and recall.

Figure 4 depicts the performance metrics in terms of precision and recall, for the test set of Dataset_1. More specifically, Model_1, which was trained from scratch performed reasonably well (93.03% precision and 92.17% recall) in the classification task. However, Model_2, which has a matching architecture with Model_1, but trained by utilizing transfer learning techniques, provides improved results, achieving 95.69% precision and 95.62% recall. Lastly, Model_3 (ResNet-50 pretrained on ImageNet dataset) results in even better outcomes (97.73% precision and 97.67% recall), due to (i) its more complex architecture, and (ii) the transfer learning capabilities, enabled by the pretrained model.

**Figure 4:**
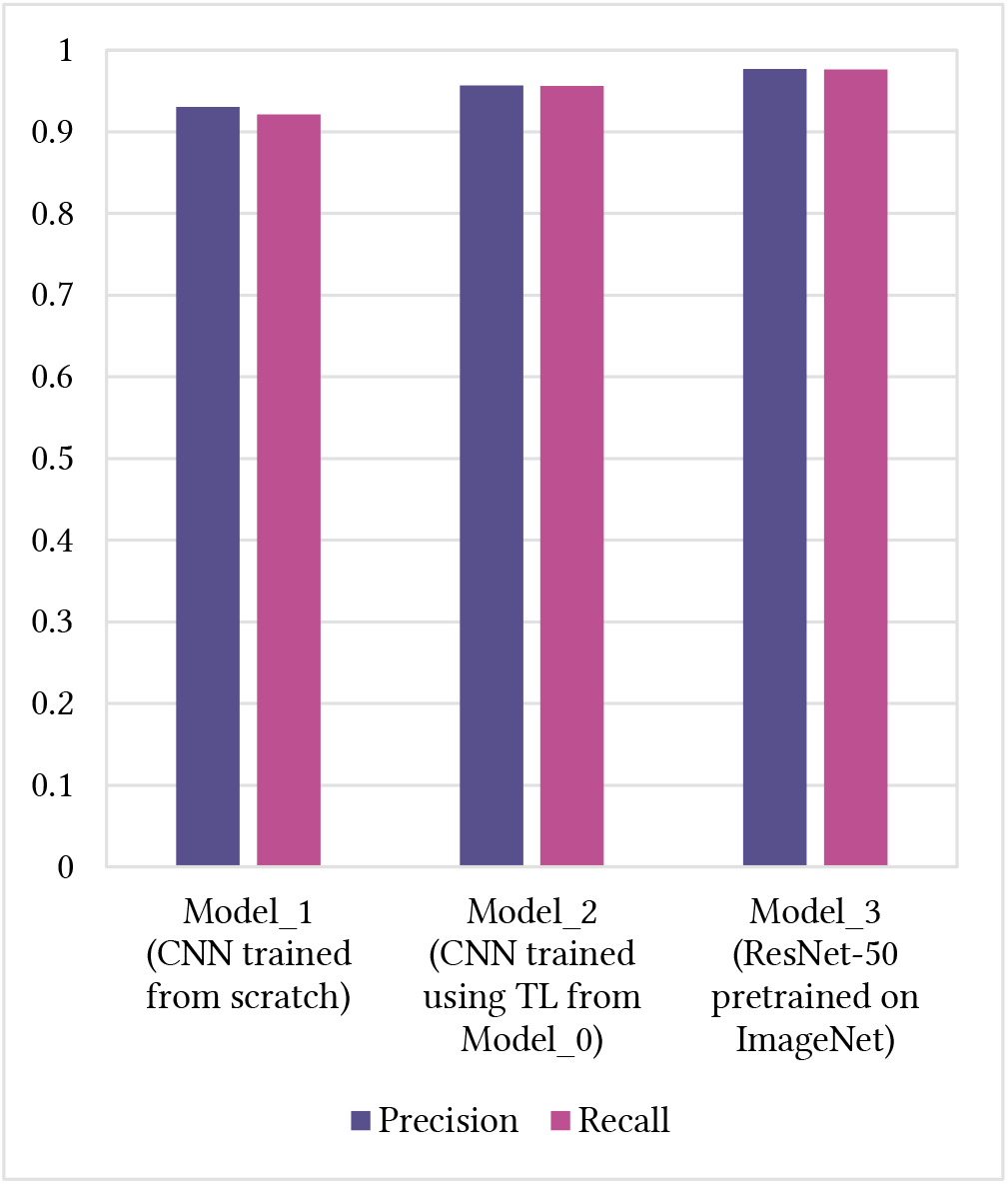
Performance metrics of the three deep models, in terms of precision and recall.

Figure 5 illustrates the performance metrics in terms of accuracy and F1-score, for the test set of Dataset_1. Similar to the above, Model_3 achieved the highest scores (97.66% accuracy and 97.61% F1-score). At the same time, Model_2 (95.65% accuracy and 95.49% F1-score) outperformed Model_1 (92.32% accuracy and 92.46% F1-score), by utilizing knowledge from other similar sources (i.e. viral pneumonia). This implies that through transfer learning the error rate of the deep learning model, which is demonstrated in Figure 3, was reduced to a factor of almost two.

**Figure 5:**
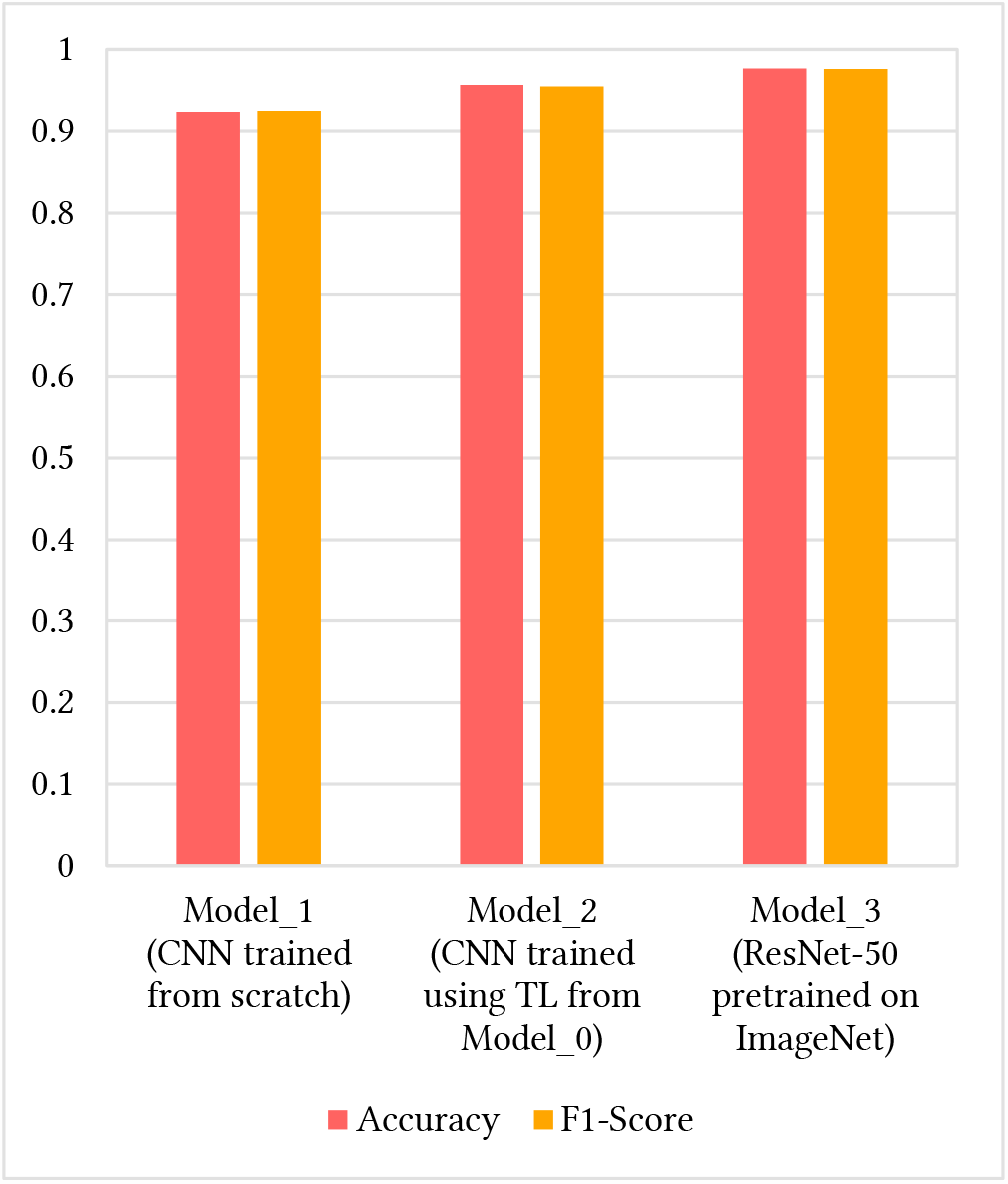
Performance metrics of the three deep models, in terms of accuracy and F1-score.

Finally, Figure 6 presents the requirements of the three classifiers, in terms of processing time. In particular, the average classification time, for an image in the test set of Dataset_1, ranges between 26.9 and 42.7 milliseconds.

**Figure 6:**
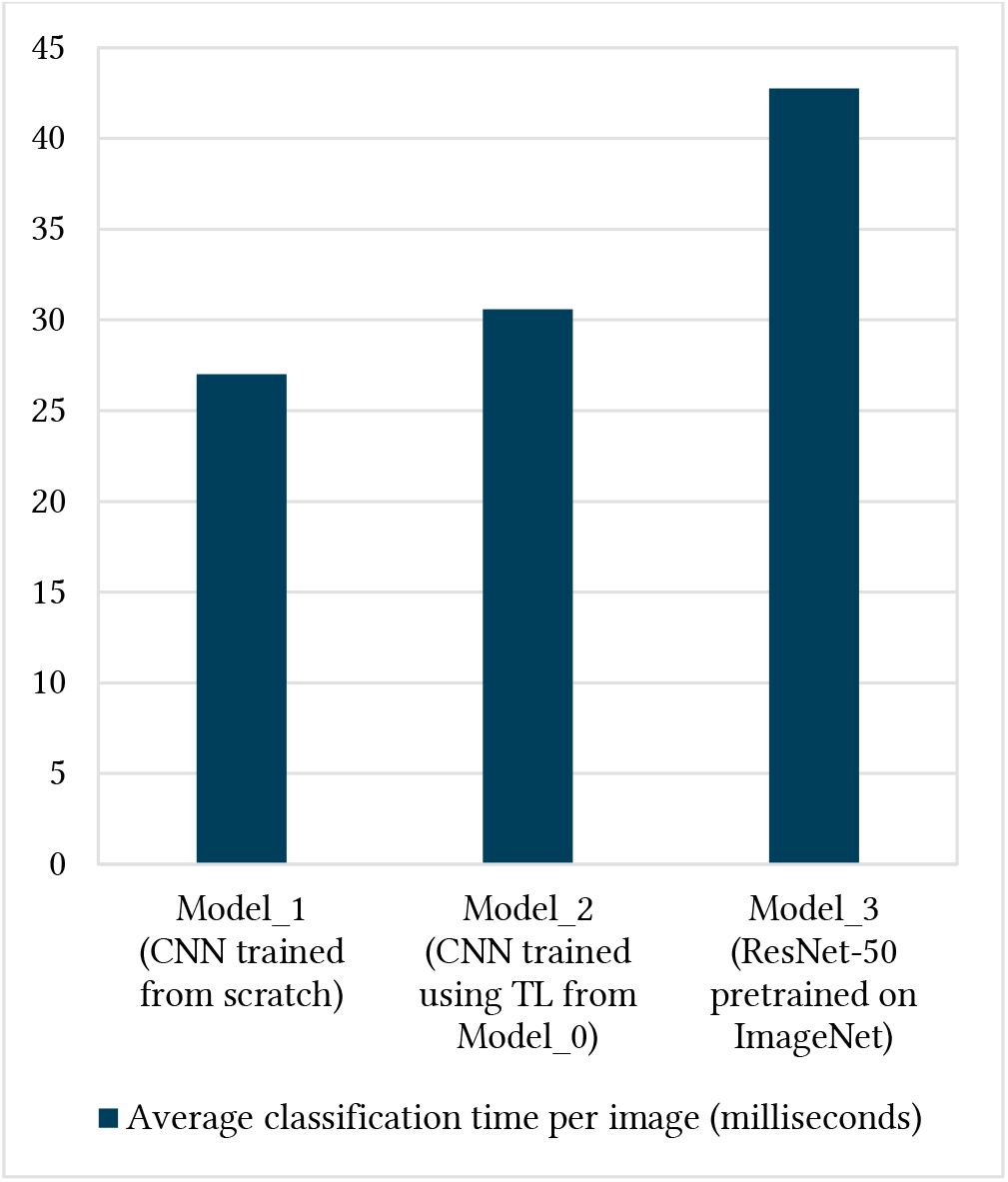
Average computational time per image, of the three deep models.

## 5 CONCLUSIONS

In this paper, we have presented a deep learning-based approach for automated detection of COVID-19 pneumonia in thoracic X-ray images. In parallel, the proposed networks differentiate COVID-19 pneumonia from bacterial pneumonia. Bearing in mind that the number of chest radiographs related to COVID-19 is limited, transfer learning can contribute to effectively train the deep models, by transferring knowledge from other data-rich sources and pretrained networks. The results of this study indicate that the transfer learning approach outperforms the performance obtained without transfer learning, for the COVID-19 classification task.

## Data Availability

The datasets used were collected from GitHub and Kaggle.

https://github.com/ieee8023/covid-chestxray-dataset

https://www.kaggle.com/paultimothymooney/chest-xray-pneumonia

## ACKNOWLEDGMENTS

This paper is supported by the H2020 Phootonics project “A Cost-Effective Photonics-based Device for Early Prediction, Monitoring and Management of Diabetic Foot Ulcers” funded under the ICT H2020 framework and the grand agreement no. 871908.

